# Multi-domain Identification of Myocardial Infarction Incidence using Explainable AI: The Overlooked Role of Periodontal Health

**DOI:** 10.64898/2025.12.18.25342537

**Authors:** Moemen Hussein, Lu Li, Karen L. Falkner, Michael J. Buck, Patricia I. Diaz, Haralampos Hatzikirou

## Abstract

Myocardial infarction (MI) is a major global health concern influenced by diverse risk factors. Despite growing evidence of oral– systemic connections, current MI models largely exclude oral health indicators, reflecting the longstanding separation between dental and medical paradigms. This study introduces a multidomain, interpretable machine learning framework that integrates detailed periodontal and oral hygiene variables, marking one of the first efforts to quantitatively incorporate these features into MI incidence identification. A population-based case-control dataset comprising 1,355 individuals and heterogeneous variables was used to train and evaluate seven supervised classifiers via nested cross-validation. Among them, XGBoost achieved the best performance (AUC = 0.88±0.01; F1 score = 0.74±0.03) and was further probability-calibrated using isotonic regression, yielding a mean Brier score of 0.14±0.01 and demonstrating well-aligned predicted probabilities. SHAP values confirmed the importance of conventional cardiovascular predictors, while several periodontal indicators such as mean clinical attachment loss, plaque index, and gingival bleeding emerged among the most influential features. Sex-stratified SHAP analysis revealed sex-specific patterns in the relative impact of oral features. Additionally, individual-level waterfall plots illustrated how oral inflammation may contribute independently or in combination with conventional factors to MI incidence identification. These findings support a systems-level view of periodontitis as a modifiable, biologically relevant factor in cardiovascular health and underscore the value of considering oral-health markers within screening and management frameworks.

## Introduction

Myocardial infarction (MI) is a major contributor to global mortality. According to the World Health Organization (WHO), cardiovascular diseases (CVDs) are the leading cause of death worldwide, accounting for approximately 32% of all annual deaths. Myocardial infarction and stroke are responsible for about 85% of these cases^1^. Although advances in acute cardiovascular care have improved short-term survival, effective MI prevention is still difficult^2,3^.

Long-used risk estimators such as the Framingham Risk Score have shaped preventive cardiology for years^4^. However, these models rely on a limited set of factors and only echo linear assumptions that may not reflect the complex, multi-factorial nature of cardiovascular risk^5,6^. In essence, these models have limited capacity to handle the high-dimensional, non-linear interactions that often characterize risk profiles^5,7^.

Machine learning (ML) is a promising tool to improve cardiovascular predictive modeling since it allows the integration of large, heterogeneous data and captures complex, non-linear associations^8,9^. However, many machine learning models operate as “black boxes,” offering little transparency about their internal logic^10^. While this lack of transparency is a major barrier to clinical use, explainable machine learning (XML) methods offer a potential solution^11,12^. Tools like Shapley Additive Explanations (SHAP) use principles of game theory to identify factors that are important at the cohort level and variables that contribute most to an individual’s outcome^13^.

Recent studies point to a strong association between emerging factors such as oral health and the likelihood of MI^14–16^. Periodontitis, in particular, has been identified as an independent risk factor for MI, even after controlling for other typical cardiovascular risk factors like age^17,18^. This link is thought to be due to induced systemic inflammation or the translocation of harmful oral bacteria through the bloodstream to other organs^19,20^. These bacteria can promote blood clotting or directly damage heart tissue and valves^19–23^. Importantly, oral hygiene practices such as brushing daily are associated with a significantly lower risk of heart-related conditions^24,25^. These associations are observed across different populations, including people with diabetes and hypertension^26,27^, highlighting the potential value of early periodontal disease detection and management in reducing MI risk and improving overall cardiovascular outcomes.

Despite these findings, oral health indicators such as periodontal metrics remain largely absent from most cardiovascular predictive models. This includes those using machine learning and deep learning, which typically rely on a mix of metabolic, clinical, lifestyle, comorbidity, and demographic data^28–35^. These oral health variables are not only inexpensive to assess but also provide early insights into chronic inflammation and behavioral patterns, representing a missed opportunity for improving MI modeling.

To address this gap, this study develops an explainable machine learning model for MI event identification that integrates clinical variables, behavioral exposures, and a rich suite of oral health indicators, including both self-reported behaviors and objective periodontal measures. We further employed SHAP to quantify the contribution of each predictor to the model’s output at both the individual and cohort levels. To our knowledge, this is among the first studies to systematically integrate both subjective and objective oral health data into an explainable machine learning framework for MI classification. By expanding the scope of cardiovascular predictive modeling to include an often-overlooked domain, our work advances the fields of preventive cardiology and precision medicine.

## Results

### Patient Characteristics

Baseline characteristics by myocardial infarction (MI) status are shown in Table 1. Among 1,355 participants, MI cases were younger on average (54.7 vs. 56.4 years; *p* < 0.001) and more likely male (76%). Differences extended to socioeconomic and oral health indicators. MI cases had fewer years of education and worse periodontal conditions, characterized by increased mean probing depth, clinical attachment loss, and plaque index (all *p* < 0.001). They also had fewer teeth and elevated bleeding on probing. Some oral hygiene practices differed significantly. Compared to controls, MI cases were less likely to floss daily or visit the dentist annually. They also reported higher tobacco consumption (23.3 vs. 10.6 pack-year) and diabetes prevalence (14.9%).

**Table 1.**
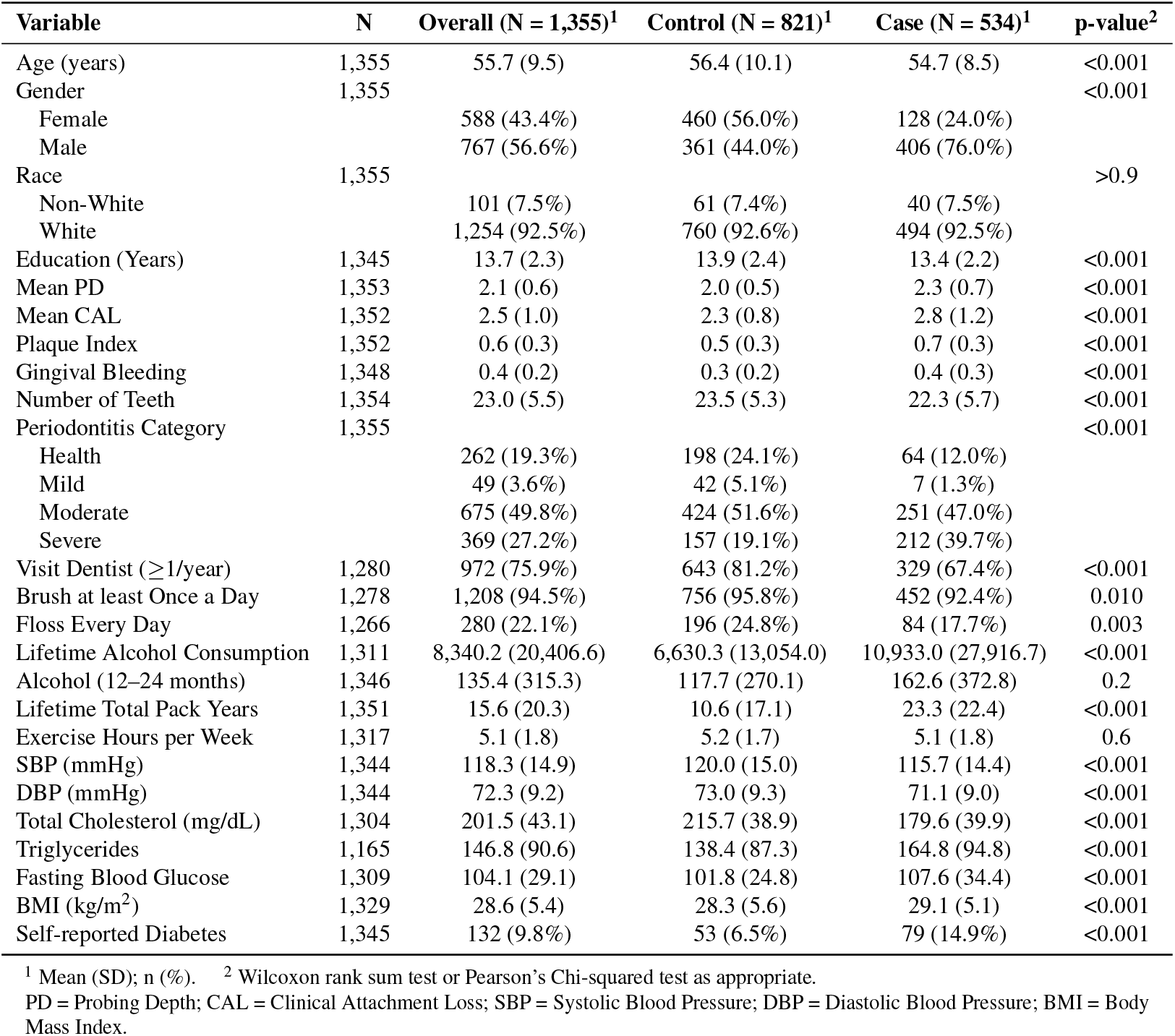
Baseline Characteristics of Study Participants Stratified by Myocardial Infarction (MI) Status.

| Variable | N | Overall (N = 1,355)<sup>1</sup> | Control (N = 821)<sup>1</sup> | Case (N = 534)<sup>1</sup> | p-value<sup>2</sup> |
| --- | --- | --- | --- | --- | --- |
| Age (years) | 1,355 | 55.7 (9.5) | 56.4 (10.1) | 54.7 (8.5) | <0.001 |
| Gender | 1,355 |  |  |  | <0.001 |
| Female |  | 588 (43.4%) | 460 (56.0%) | 128 (24.0%) |  |
| Male |  | 767 (56.6%) | 361 (44.0%) | 406 (76.0%) |  |
| Race | 1,355 |  |  |  | >0.9 |
| Non-White |  | 101 (7.5%) | 61 (7.4%) | 40 (7.5%) |  |
| White |  | 1,254 (92.5%) | 760 (92.6%) | 494 (92.5%) |  |
| Education (Years) | 1,345 | 13.7 (2.3) | 13.9 (2.4) | 13.4 (2.2) | <0.001 |
| Mean PD | 1,353 | 2.1 (0.6) | 2.0 (0.5) | 2.3 (0.7) | <0.001 |
| Mean CAL | 1,352 | 2.5 (1.0) | 2.3 (0.8) | 2.8 (1.2) | <0.001 |
| Plaque Index | 1,352 | 0.6 (0.3) | 0.5 (0.3) | 0.7 (0.3) | <0.001 |
| Gingival Bleeding | 1,348 | 0.4 (0.2) | 0.3 (0.2) | 0.4 (0.3) | <0.001 |
| Number of Teeth | 1,354 | 23.0 (5.5) | 23.5 (5.3) | 22.3 (5.7) | <0.001 |
| Periodontitis Category | 1,355 |  |  |  | <0.001 |
| Health |  | 262 (19.3%) | 198 (24.1%) | 64 (12.0%) |  |
| Mild |  | 49 (3.6%) | 42 (5.1%) | 7 (1.3%) |  |
| Moderate |  | 675 (49.8%) | 424 (51.6%) | 251 (47.0%) |  |
| Severe |  | 369 (27.2%) | 157 (19.1%) | 212 (39.7%) |  |
| Visit Dentist (≥ 1/year) | 1,280 | 972 (75.9%) | 643 (81.2%) | 329 (67.4%) | <0.001 |
| Brush at least Once a Day | 1,278 | 1,208 (94.5%) | 756 (95.8%) | 452 (92.4%) | 0.010 |
| Floss Every Day | 1,266 | 280 (22.1%) | 196 (24.8%) | 84 (17.7%) | 0.003 |
| Lifetime Alcohol Consumption | 1,311 | 8,340.2 (20,406.6) | 6,630.3 (13,054.0) | 10,933.0 (27,916.7) | <0.001 |
| Alcohol (12–24 months) | 1,346 | 135.4 (315.3) | 117.7 (270.1) | 162.6 (372.8) | 0.2 |
| Lifetime Total Pack Years | 1,351 | 15.6 (20.3) | 10.6 (17.1) | 23.3 (22.4) | <0.001 |
| Exercise Hours per Week | 1,317 | 5.1 (1.8) | 5.2 (1.7) | 5.1 (1.8) | 0.6 |
| SBP (mmHg) | 1,344 | 118.3 (14.9) | 120.0 (15.0) | 115.7 (14.4) | <0.001 |
| DBP (mmHg) | 1,344 | 72.3 (9.2) | 73.0 (9.3) | 71.1 (9.0) | <0.001 |
| Total Cholesterol (mg/dL) | 1,304 | 201.5 (43.1) | 215.7 (38.9) | 179.6 (39.9) | <0.001 |
| Triglycerides | 1,165 | 146.8 (90.6) | 138.4 (87.3) | 164.8 (94.8) | <0.001 |
| Fasting Blood Glucose | 1,309 | 104.1 (29.1) | 101.8 (24.8) | 107.6 (34.4) | <0.001 |
| BMI (kg/m<sup>2</sup>) | 1,329 | 28.6 (5.4) | 28.3 (5.6) | 29.1 (5.1) | <0.001 |
| Self-reported Diabetes | 1,345 | 132 (9.8%) | 53 (6.5%) | 79 (14.9%) | <0.001 |

<sup>1</sup> Mean (SD); n (%). <sup>2</sup> Wilcoxon rank sum test or Pearson's Chi-squared test as appropriate.
PD = Probing Depth; CAL = Clinical Attachment Loss; SBP = Systolic Blood Pressure; DBP = Diastolic Blood Pressure; BMI = Body Mass Index.

### Model Development and Evaluation

We trained and evaluated seven ML classifiers using a nested cross-validation framework (see Fig. 1). This approach ensured an unbiased assessment of model performance and allowed systematic hyperparameter optimization.

**Figure 1.**
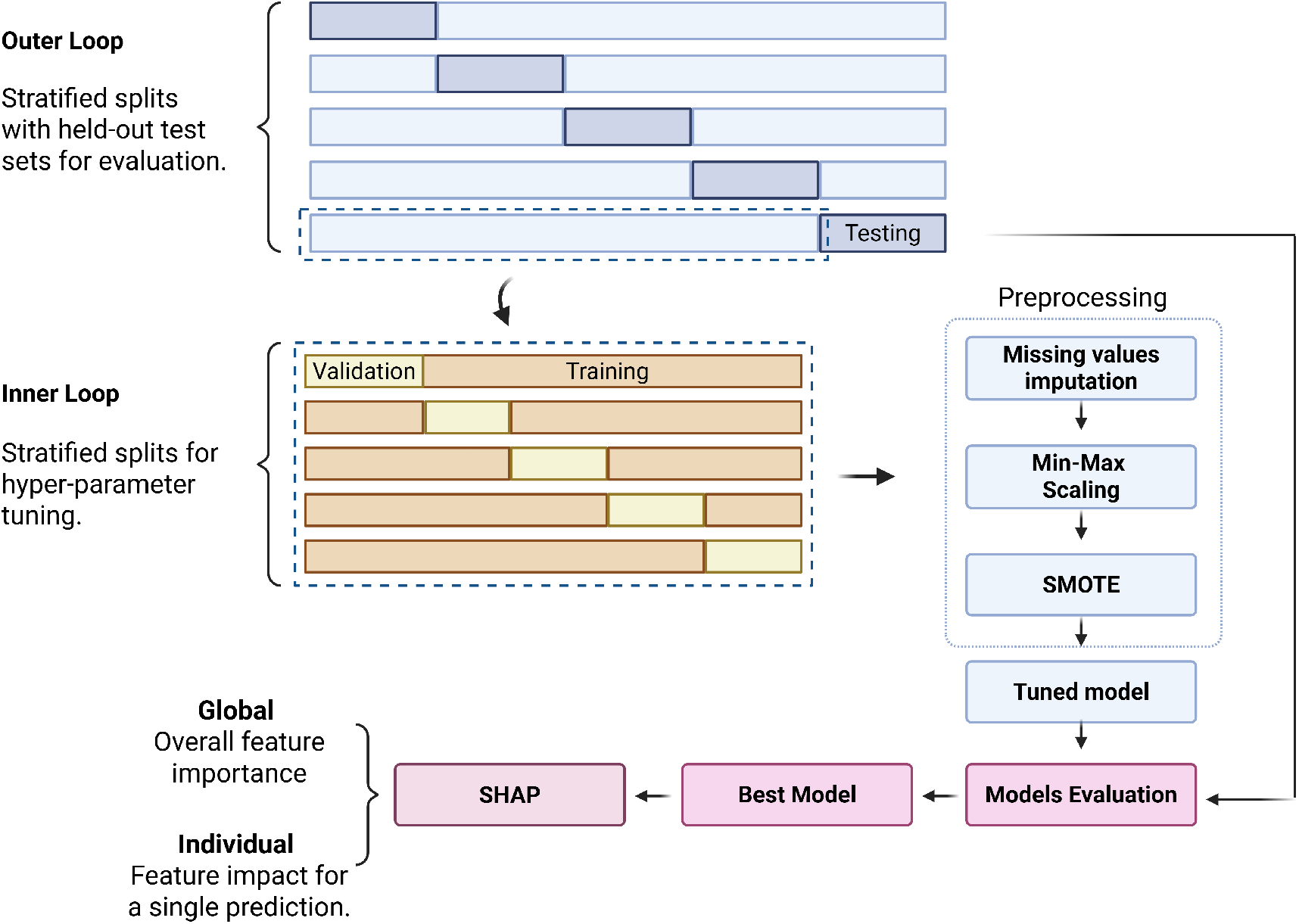
Schematic of nested Cross-Validation framework for model training and evaluation. The outer loop consists of stratified splits with held-out test sets used solely for final model evaluation. Within each outer fold, an inner loop performs stratified cross-validation to optimize hyperparameters. Preprocessing steps including missing value imputation, Min-Max scaling, and class imbalance handling via SMOTE are applied exclusively within the training folds to prevent information leakage. The resulting tuned models are evaluated and compared across outer folds. The best-performing model is then interpreted using SHAP analysis, providing both global feature importance and individual prediction-level insights.

#### Hyperparameter Optimization

The hyperparameter search spaces and corresponding optimal values for each model, identified via Bayesian optimization within the nested cross-validation framework, are presented in Table 2. XGBoost, the best-performing model, was tuned for high complexity. Using many trees (285) with deep structures (max_depth = 9), it is capable of learning detailed patterns in the data. High values for the subsample (0.94) and colsample_bytree (0.91) parameters allowed each tree to use most of the training data and features. A relatively low learning rate (0.03) improves training stability, while a higher split threshold (gamma = 0.86) reduces overfitting by requiring high gain for each split.

**Table 2.** Hyperparameter Search Spaces and Optimization Results for Machine Learning Models.

| Model | Parameter | Search Space<sup>1,2</sup> | Best Value |
| --- | --- | --- | --- |
| XGBoost | n_estimators | 50–500 | 285 |
|  | max_depth | 3–10 | 9 |
| | learning_rate | $10^{-3}$–0.3 (log-uniform) | 0.030 |
|  | subsample | 0.5–1.0 | 0.94 |
|  | colsample_bytree | 0.5–1.0 | 0.91 |
|  | gamma | 0–5 | 0.86 |
| | reg_alpha | $10^{-8}$–10 (log-uniform) | $7.23 \times 10^{-8}$ |
| | reg_lambda | $10^{-8}$–10 (log-uniform) | 0.063 |
| Random Forest | n_estimators | 50–500 | 120 |
|  | max_depth | 5–30 | 8 |
|  | min_samples_split | 2–20 | 10 |
|  | min_samples_leaf | 1–20 | 5 |
|  | max_features | 0.1–1.0 | 0.49 |
|  | criterion | {gini, entropy} | entropy |
|  | bootstrap | {True, False} | True |
| Decision Tree | max_depth | 1–30 | 25 |
|  | min_samples_split | 2–50 | 30 |
|  | min_samples_leaf | 1–30 | 8 |
|  | max_features | {sqrt, log2, None} | None |
|  | criterion | {gini, entropy} | gini |
| KNN | n_neighbors | 1–50 | 13 |
|  | weights | {uniform, distance} | distance |
|  | p | {1, 2} | 2 |
|  | metric | {minkowski, euclidean, manhattan} | manhattan |
| Naive Bayes | var_smoothing | $10^{-10}$–$10^{-3}$ (log-uniform) | 0.00049 |
| Log. Regression | C | $10^{-4}$–$10^2$ (log-uniform) | 1.73 |
|  | penalty | {l1, l2} | l1 |
|  | solver | {liblinear, saga} | liblinear |
| SVC | C | $10^{-3}$–$10^3$ (log-uniform) | 931.43 |
|  | kernel | {linear, rbf, poly, sigmoid} | rbf |
| | gamma | $10^{-4}$–$10^1$ (log-uniform) | 0.009 |
|  | degree | 2–5 | 3 |

All parameters were tuned using Bayesian optimization with cross-validation.
<sup>1</sup>Categorical search spaces are denoted with braces {...}.
<sup>2</sup>Log-uniform indicates sampling was performed in log-space to capture values across several orders of magnitude.

#### Model Performance

The performance summary in Table 3 indicates the advantage of ensemble learning methods for this classification task. Based on results from five different testing folds, XGBoost emerged as the best algorithm according to most metrics. By achieving the highest mean AUC (0.88 ± 0.01) and F1 score (0.74 ± 0.03), it reflected strong overall classification performance and a good balance between sensitivity and specificity. As shown in Figure 2, the Receiver Operating Characteristic (ROC) curve for each testing fold illustrates the model’s robust performance. This strong performance was further supported by the Matthews Correlation Coefficient (MCC) value (0.58 ± 0.04), indicating a high correlation between the true and predicted classes. Random Forest closely followed XGBoost but showed slightly lower performance values and a higher standard deviation (MCC: 0.56 ± 0.05).

**Table 3.**
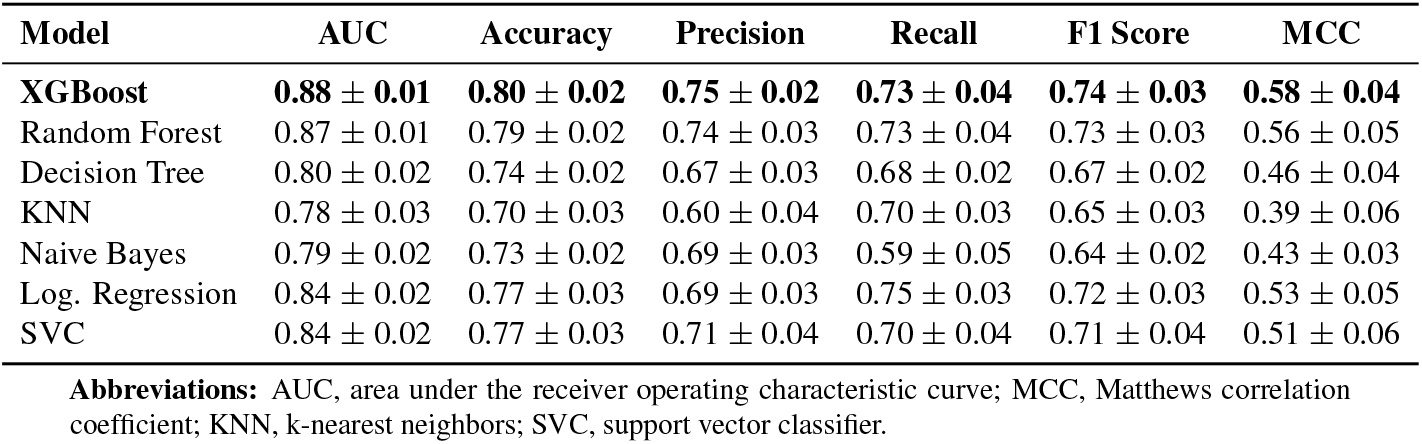
Performance of Machine Learning Models for Myocardial Infarction Prediction.

**Figure 2.**
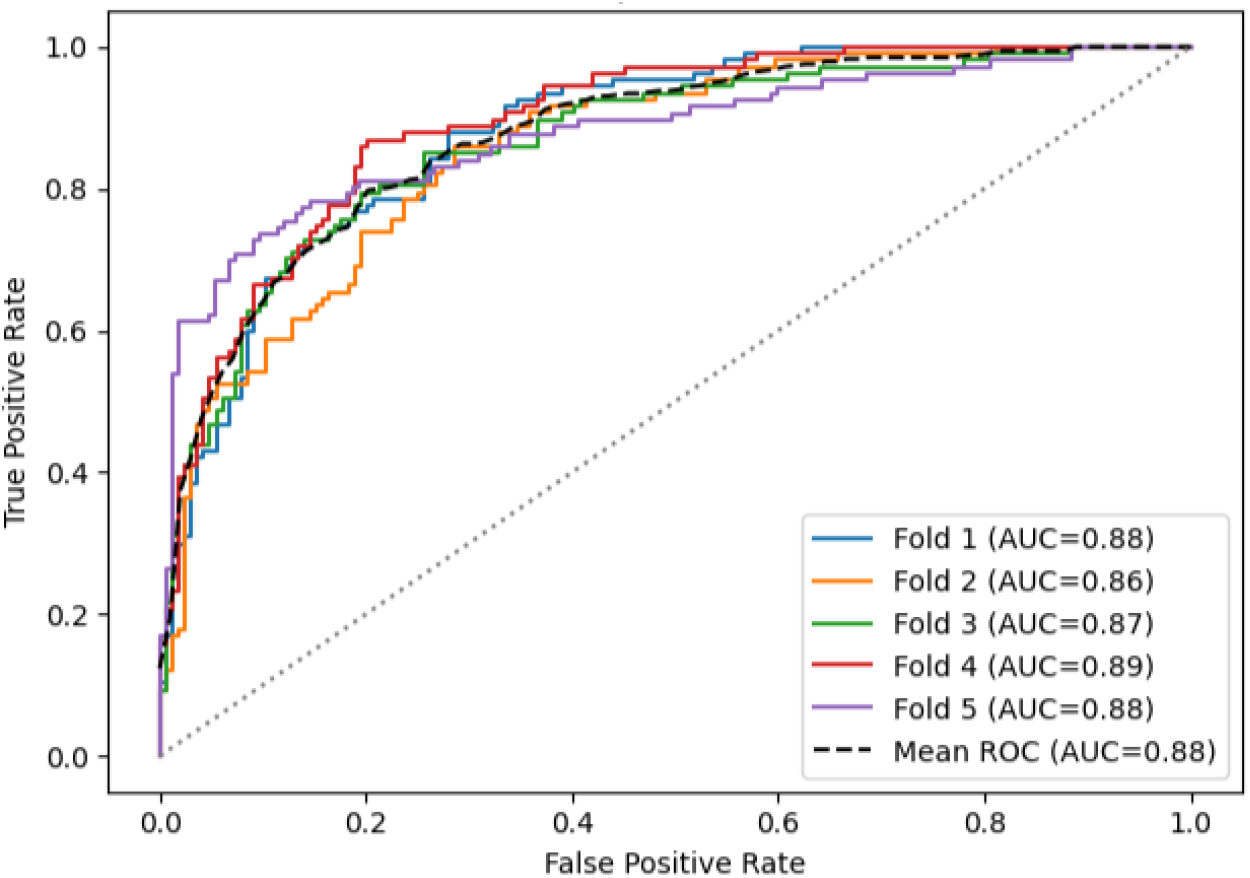
Receiver operating characteristic (ROC) curves from five outer folds of a nested cross-validation framework demonstrate consistently high discriminative performance (mean AUC = 0.88). The tight clustering of curves underscores the model’s reliability across testing sets.

Support Vector Machine and Logistic Regression models ranked next after the ensemble models with identical AUCs of 0.84 ± 0.02 but slightly different F1 and MCC values. The Decision Tree model followed, showing moderate performance with an AUC of 0.80 and an F1 score of 0.67. K-Nearest Neighbors and Naive Bayes ranked lowest, with lower AUC values (0.78–0.79) and F1 scores (0.64–0.65), reflecting limited capacity to capture complex relationships.

#### Calibration Assessment

To account for predictive uncertainty, model calibration is important to ensure that predicted probabilities align with the true frequencies of outcomes. This enhances the reliability of the results and supports better decision making. Here, we applied isotonic regression to adjust the best-performing model (XGBoost) outputs. As can be seen in Figure 3, the mean calibration curve closely followed the diagonal, and the smoothed 95% confidence band that shows the fold-to-fold variability remained narrow throughout most of the probability range. From a qualitative perspective, the classifier showed strong calibration with a mean Brier score of 0.14 ± 0.009 across the testing sets, indicating that the predicted probabilities closely matched the observed outcomes.

**Figure 3.**
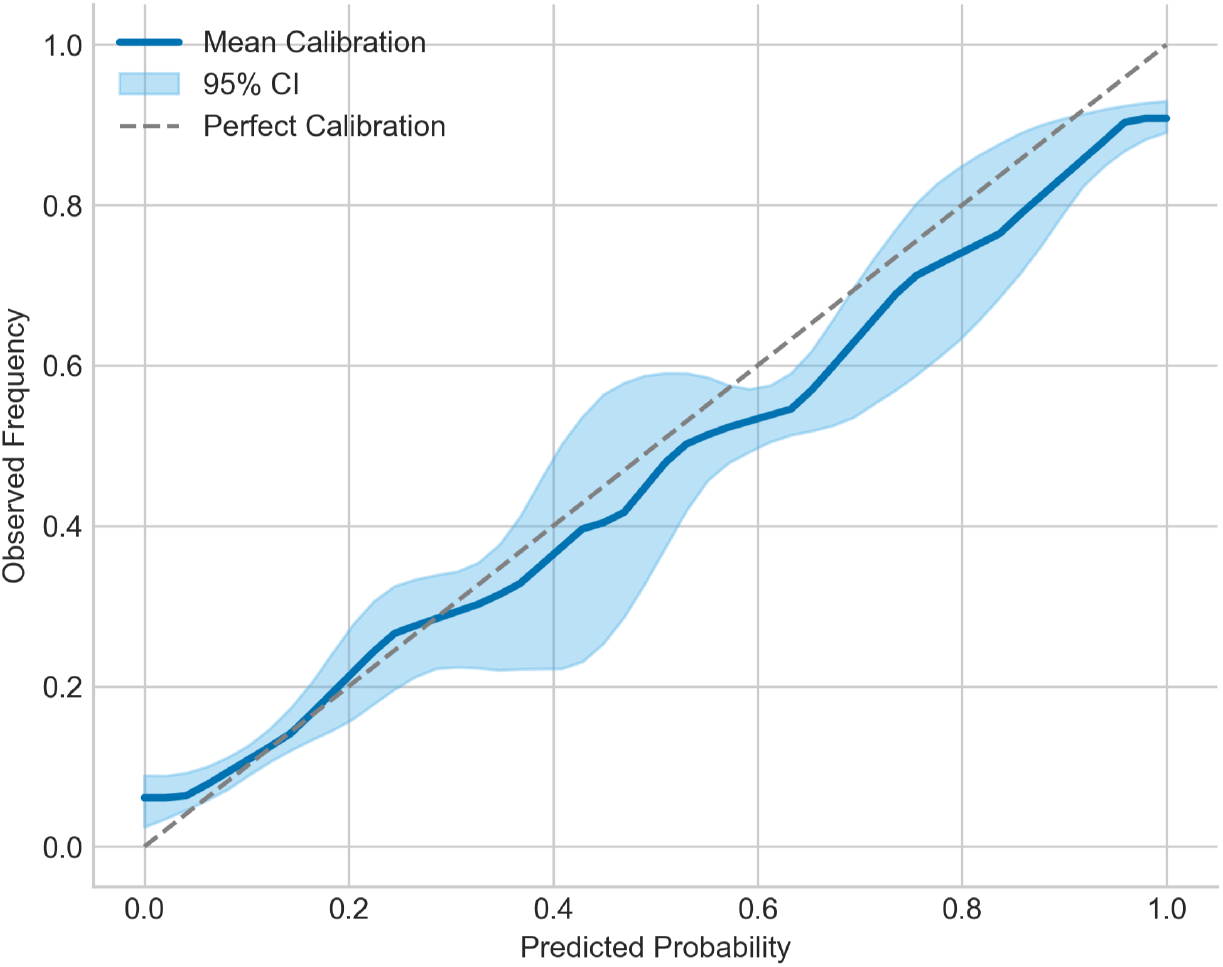
Calibration curve with a smoothed 95% confidence band for the isotonic regression calibrated XGBoost model. The mean calibration curve (solid blue line) closely follows the diagonal (dashed gray line), indicating that predicted probabilities align well with observed outcome frequencies. The narrow 95% confidence band (shaded area) across cross-validation folds reflects low variability and good calibration stability.

### Model Interpretation

We applied SHapley Additive exPlanations (SHAP) to quantify the contribution of each feature to predictions made by the best model (XGBoost). SHAP is built on cooperative game theory to ensure fair allocation of a model’s prediction among its input features. For example, it assigns zero credit to features that do not influence the model (dummy axiom) and follows the symmetry principle when two features behave identically. This analysis was conducted at both the global and individual levels, enabling detailed assessment of the predictive importance of oral health indicators such as mean probing depth, clinical attachment loss, gingival bleeding, and plaque index.

#### Global Feature Importance

Figure 4 presents the SHAP summary bar plot, which ranks features based on their mean absolute SHAP values, reflecting their overall impact on the model’s output. As expected, traditional cardiovascular risk factors such as total cholesterol and triglyceride levels emerged as the most important predictors. Notably, several indicators of periodontal health also ranked high, including mean clinical attachment loss (CAL), plaque index, number of teeth, and gingival bleeding, highlighting their potential in cardiovascular predictive modeling. This underscores the value of using diverse features to model complex conditions like MI.

**Figure 4.**
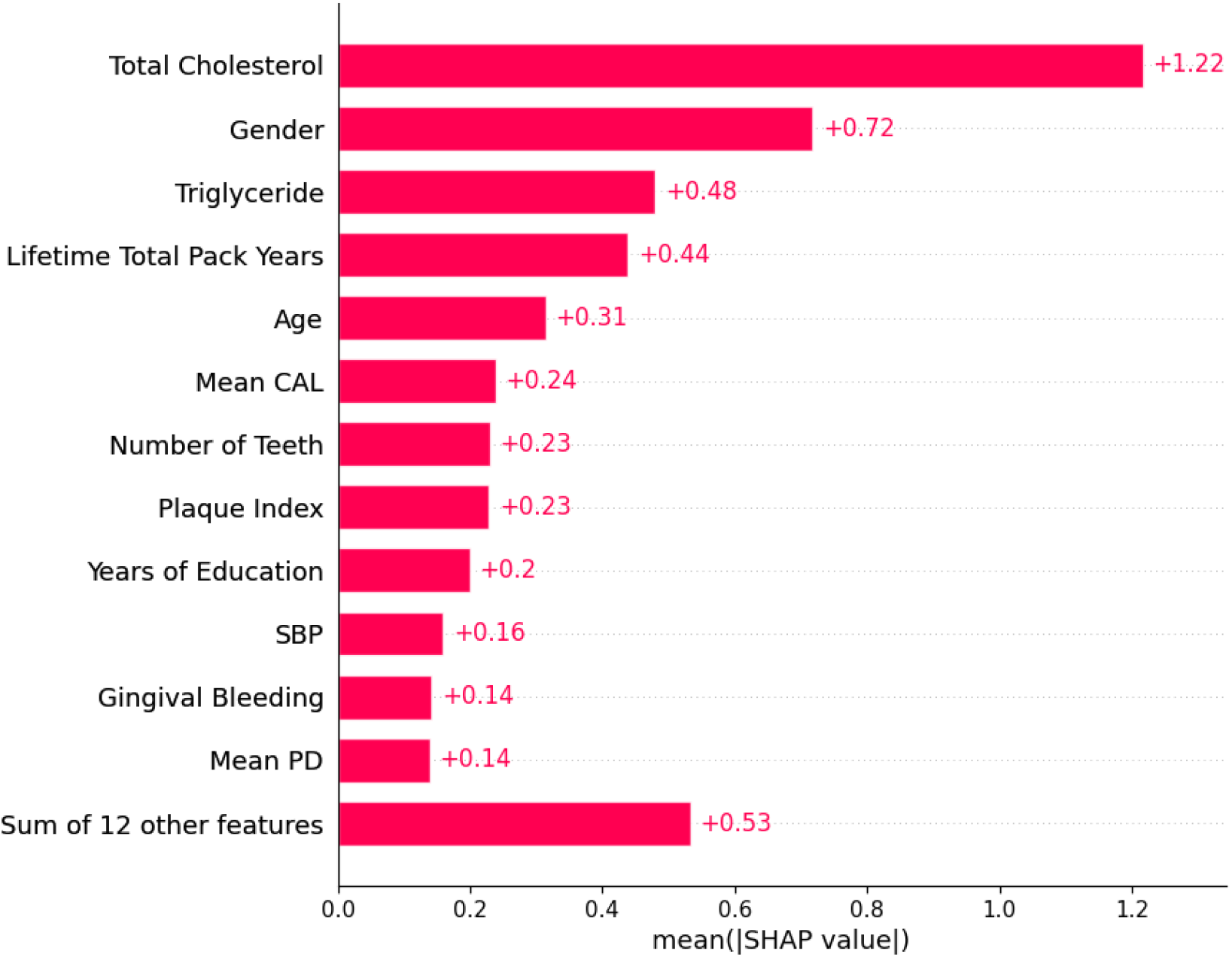
Global-level feature importance analysis. SHAP summary plot for the best-performing model (XGBoost), showing the mean absolute SHAP value for each predictor, reflecting overall influence on myocardial infarction risk classification.

#### Sex-Specific Feature Importance

To explore potential sex-specific patterns in feature importance, we performed a stratified SHAP analysis by biological sex. As shown in Figure 5, the bar plot illustrates the mean absolute SHAP values for male (*n* = 767) and female (*n* = 588) participants, allowing direct comparison across groups. Although total cholesterol was the top predictor in both sexes, it had a greater impact in males (1.35) compared to females (1.04), suggesting sex-based differences in its role in disease. Interestingly, the model showed differences between males and females when using gender as a predictive feature. This may reflect complex interactions with other covariates, such as lifestyle behaviors, embedded in the data. Age also appeared more influential in males (0.39 vs. 0.22), which may indicate earlier or more aggressive MI progression. Conversely, oral features such as plaque index (0.28 vs. 0.19) and number of teeth (0.27 vs. 0.20) had greater influence in females, suggesting a more pronounced role for oral health in cardiovascular risk among women.

**Figure 5.**
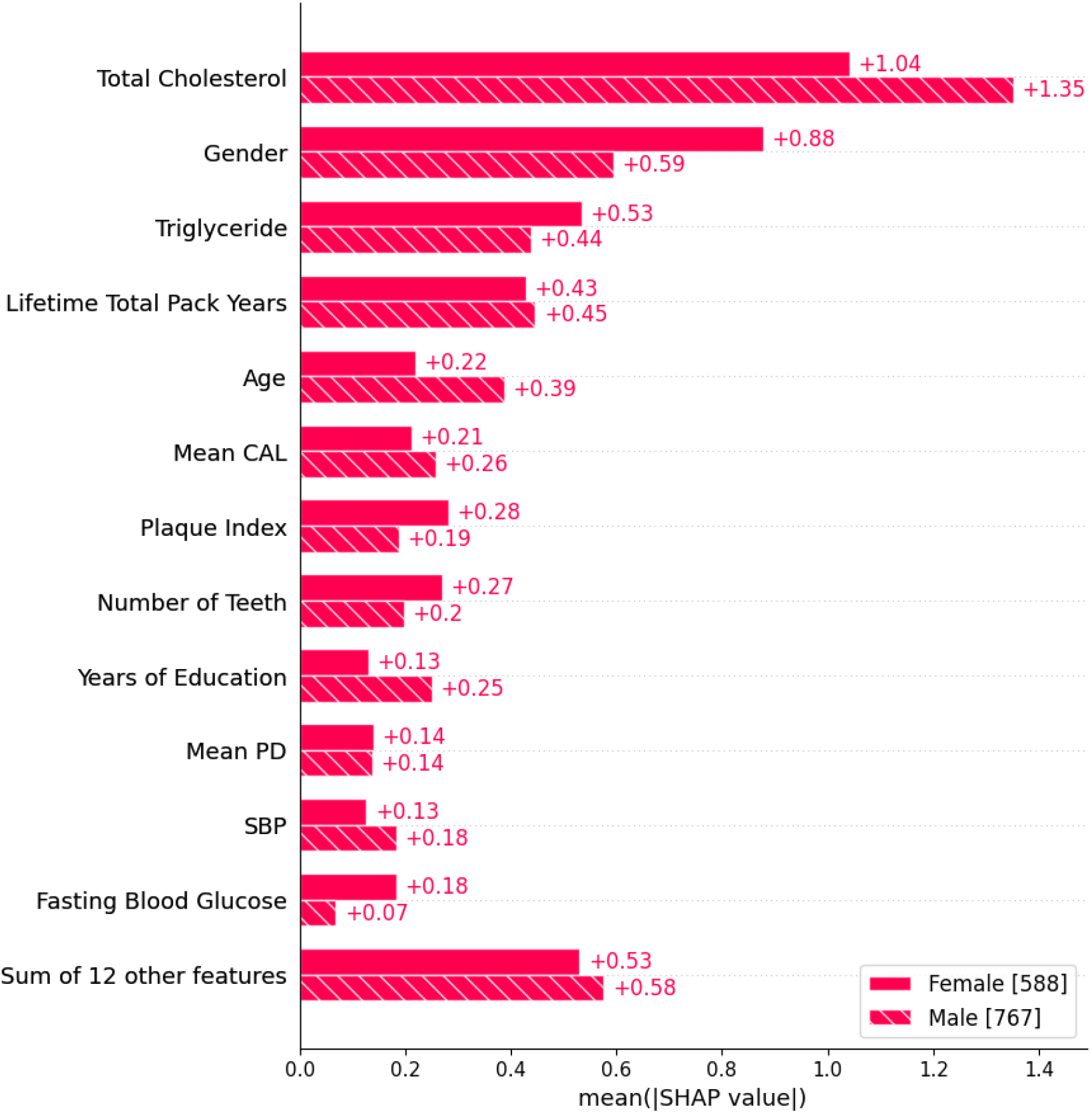
Sex-Stratified SHAP analysis of feature importance. Bar plots compare mean absolute SHAP values for key predictors in male and female participants, revealing sex-specific differences in risk attribution for myocardial infarction.

#### Periodontitis Severity and Predictive Contribution

To assess the impact of periodontitis severity on model predictions, we compared SHAP values for the categorical periodontitis status variable. As shown in Figure 6, SHAP values increased with greater periodontitis severity, indicating a monotonic trend and suggesting a dose-response relationship. To statistically validate this pattern, we conducted pairwise comparisons using the Mann-Whitney test with Bonferroni correction. Results confirmed that SHAP values differed significantly between all periodontitis categories (*p* < 0.001), indicating that each increase in periodontal severity corresponded to a statistically significant rise in its contribution to MI incidence identification.

**Figure 6.**
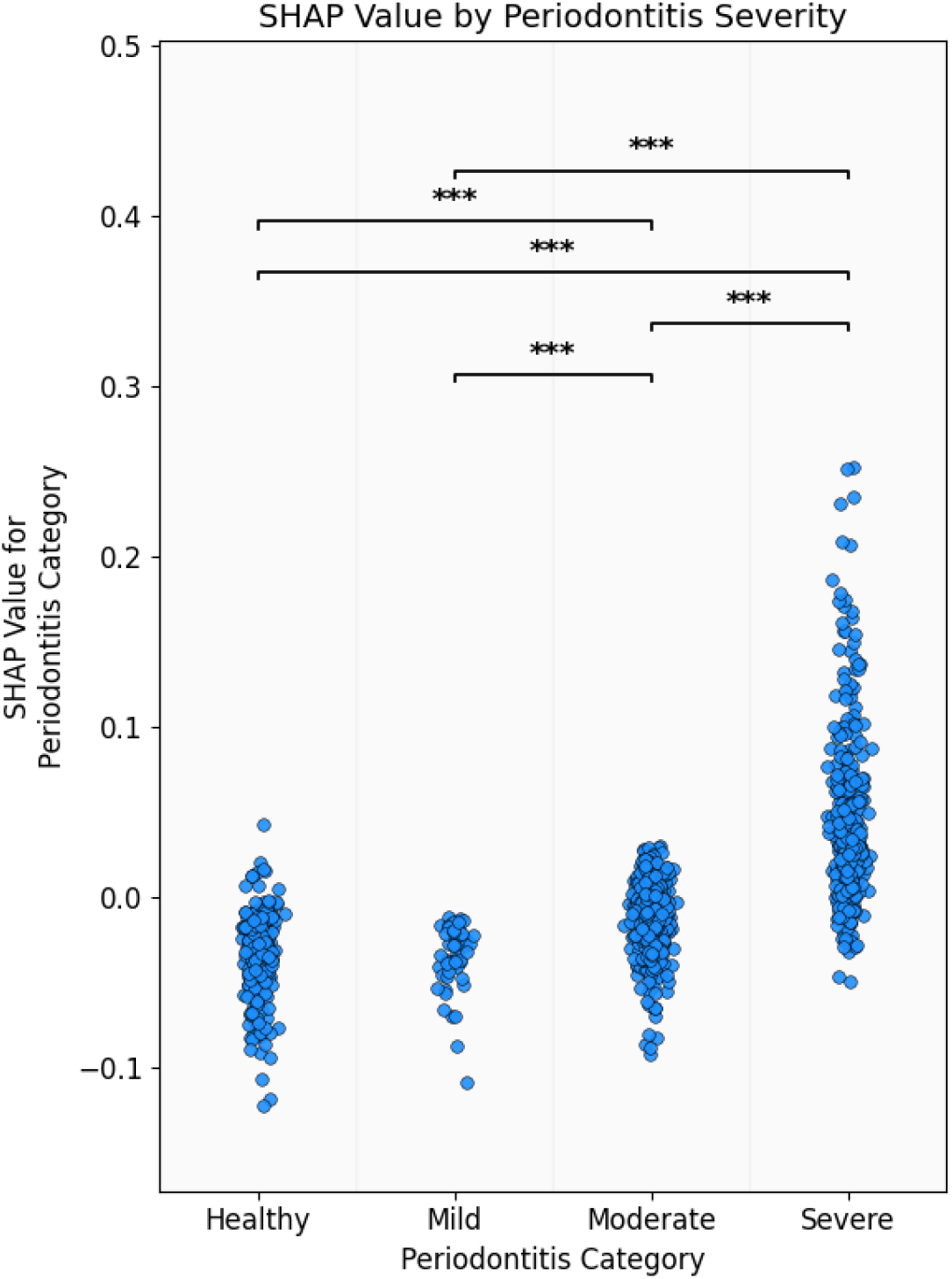
Distributions of SHAP values across periodontitis categories show that feature importance shifts with disease progression. Statistically significant pairwise differences, tested via Mann–Whitney U. Significant differences (*p* < 0.001) were visualized using bracket annotations.

#### Instance-Level Interpretability

To complement global and group-level findings, we used SHAP waterfall plots to showcase individual-level predictions for two participants (Figures 7, 8). These plots decompose a single prediction by showing how each feature contributes to the outcome, starting from a reference expected value. Red bars indicate positive shifts (increased likelihood), while blue bars indicate negative shifts (decreased likelihood). The final prediction is the sum of all feature contributions.

**Figure 7.**
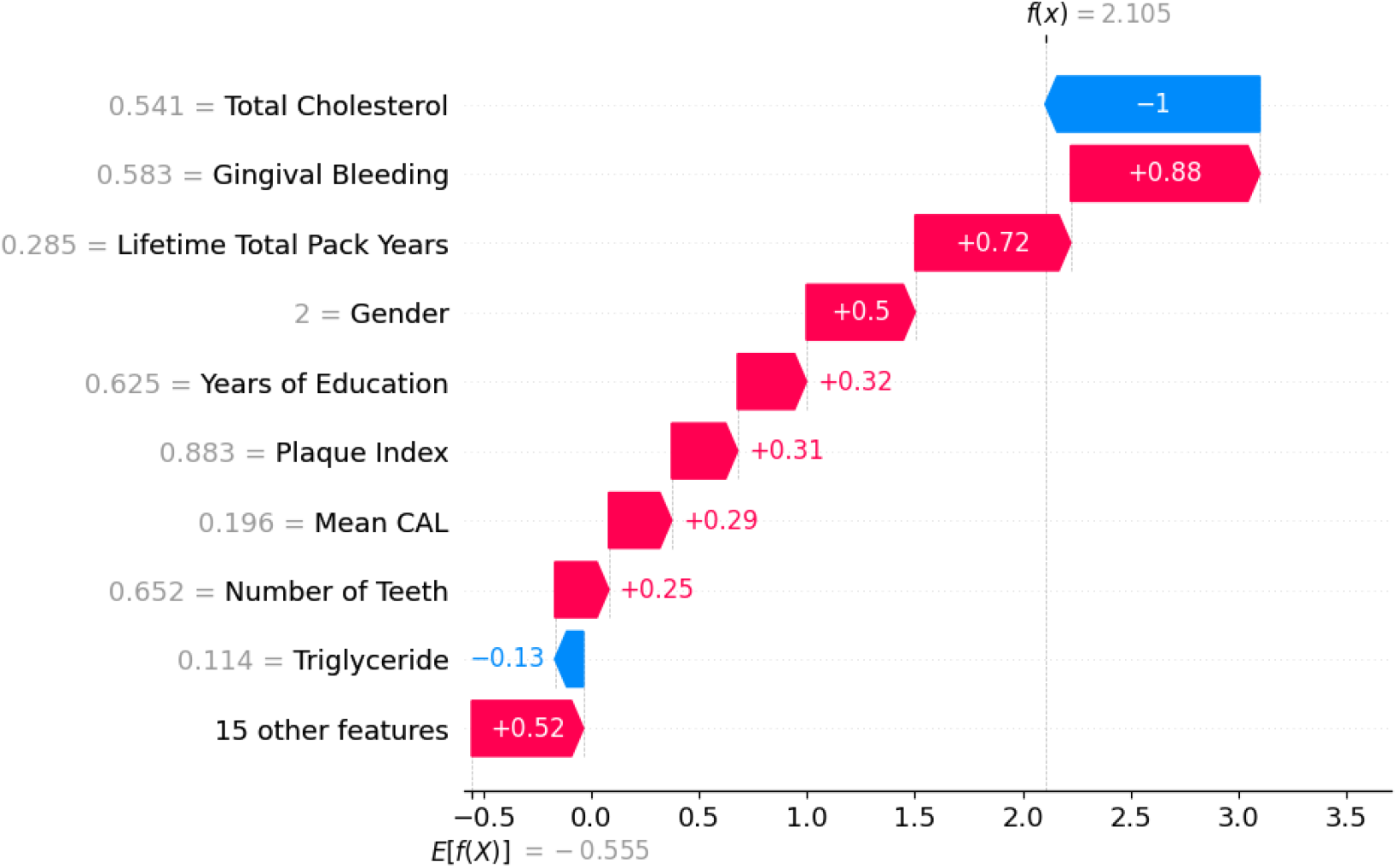
Individual-level interpretability for a high-risk prediction (f(x) = 2.105). In this case, MI risk was primarily driven by gingival bleeding, tobacco exposure, and male sex. Notably, total cholesterol contributed a protective signal, an effect likely influenced by medication use. This highlights the complex interplay of predictors in model decision-making.

**Figure 8.**
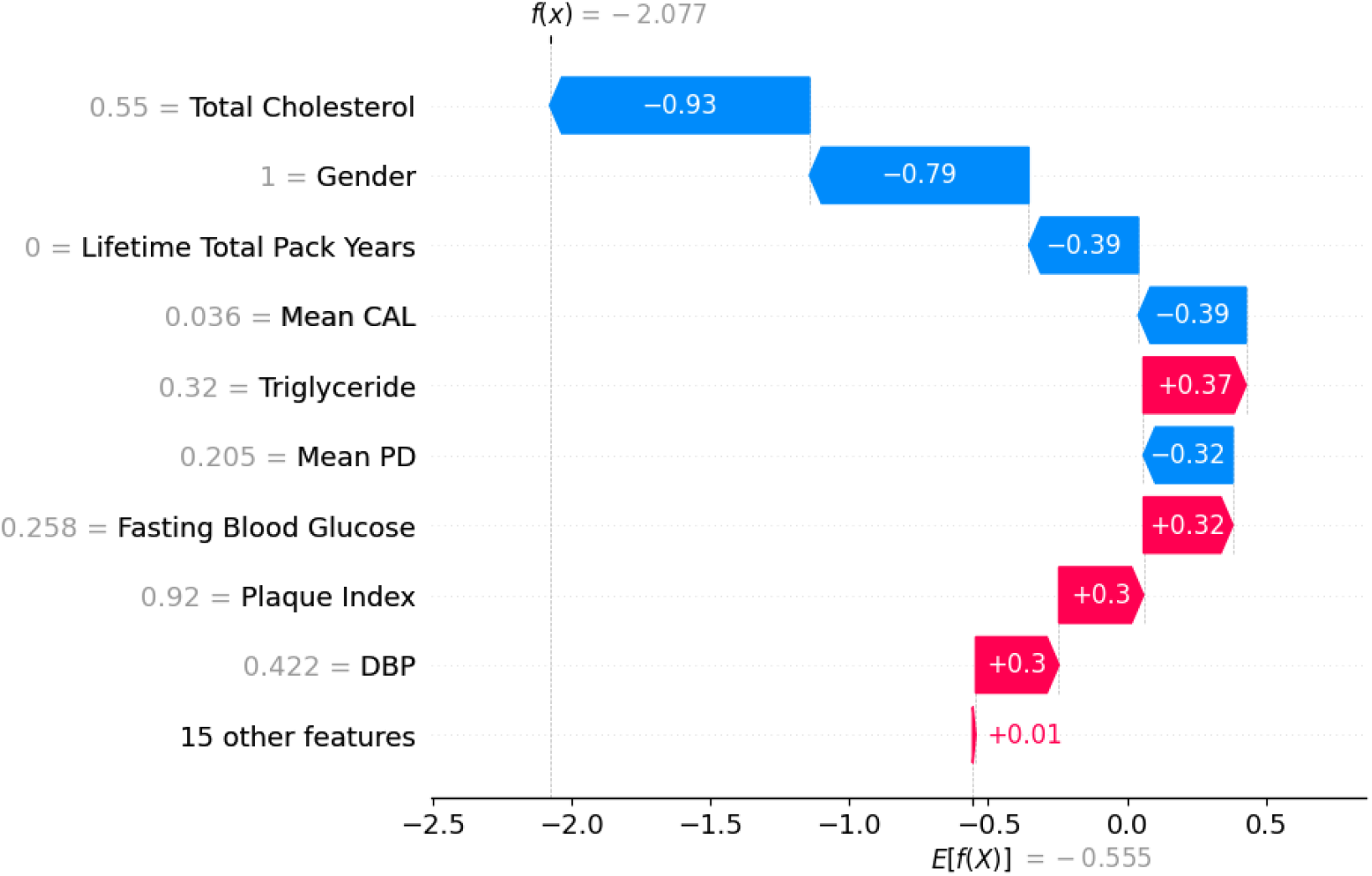
Protective predictors in a low-risk profile (f(x) = –2.077). A low-risk prediction demonstrates dominant protective effects from elevated Total Cholesterol and female sex. Subtle risk signals were contributed by triglyceride levels and periodontal indicators, highlighting multifactorial buffering against MI.

Figure 7 shows a high-likelihood prediction ( *f* (*x*) = 2.105), largely driven by elevated gingival bleeding, smoking, lower education, and male sex. Although total cholesterol appeared to have a strong protective effect, it could not offset the cumulative impact of pro-inflammatory and behavioral factors, especially periodontal indicators like CAL and plaque index. The apparent protective effect of total cholesterol likely reflects statin use, highlighting the limitations of relying on isolated features in the management of complex, multifactorial conditions like MI.

In contrast, Figure 8 presents a low-likelihood prediction ( *f* (*x*) = −2.077), where total cholesterol and female sex were the main protective factors. Additional protective contributions came from no smoking history, low CAL, and shallow probing depth. While features like triglycerides, fasting glucose, and plaque index slightly increased the predicted likelihood, their effects were outweighed by stronger protective factors.

## Discussion

This study introduces an interpretable machine learning framework that integrates heterogeneous data to identify myocardial infarction (MI) incidence. While machine learning is increasingly used for cardiovascular modeling, this work stands out for expanding the feature space to include periodontal and oral hygiene variables and for applying explainable techniques to understand their contribution to the prediction. The findings are significant not only for improving predictive performance but also for shedding light on previously underexplored predictive links between oral and cardiovascular health, offering insight into the systemic nature of oral health.

Mechanistically, the potential role of periodontal indicators in MI prediction is supported by increasing evidence. Chronic periodontitis is a long-lasting infection that promotes low-grade systemic inflammation. Such inflammation has been associated with vascular damage through pro-inflammatory cytokines like IL-1*β*, IL-6, and TNF-*α*^19, 36^. These cytokines are known to drive endothelial dysfunction, monocyte adhesion, and smooth muscle proliferation, which are key steps in atherogenesis^19,37^. In addition, pathogen-associated molecular patterns (PAMPs) from oral bacteria such as *Porphyromonas gingivalis* and *Aggregatibacter actinomycetemcomitans* have been found in atherosclerotic plaques and shown the ability to speed up lipid deposition and blood vessel inflammation^19,37^. These molecular mechanisms suggest a pathway by which oral diseases contribute to cardiovascular issues. The demonstrated predictive value of features like clinical attachment loss, gingival bleeding, and plaque index supports the view that oral disease markers may reflect systemic inflammatory burden relevant to cardiovascular risk.

From a modeling perspective, the superior performance of ensemble models, e.g., XGBoost, points to the importance of nonlinear interactions in MI identification. Linear regression models assume additive effects, potentially missing threshold dynamics and higher-order interactions. Our results show that oral variables such as CAL and plaque index interact in different ways with systemic measures like total cholesterol and fasting glucose. These interactions lead to personalized risk profiles that linear models may overlook. For example, in some cases, severe periodontal inflammation canceled out the protective effect of high total cholesterol. In others, periodontal measures signaled low to mild predicted likelihood, aligning with other systemic indicators.

The use of SHapley Additive exPlanations (SHAP) enabled transparent interpretation of model outputs. At the cohort level, SHAP summary plots ranked periodontal variables among the top predictors, outperforming some widely used markers like fasting glucose or blood pressure. SHAP waterfall plots showed that different features influenced the prediction depending on the individual. Some features, such as smoking, being male, and elevated plaque or CAL, increased likelihood, while others, including high total cholesterol and low gum inflammation, lowered it. These plots provided an understanding of feature contributions that could guide patient or clinician decisions.

Sex-stratified analyses revealed additional heterogeneity in model attribution. While total cholesterol was still the most influential in both sexes, the relative importance varied, with a stronger effect found in males. A similar pattern was observed for age and CAL. Conversely, other variables like dietary fats and plaque index had greater impact in women. Such patterns may reflect biological factors (e.g., sex hormones, immune response) as well as behavioral and social determinants like healthcare access or oral hygiene habits.

Due to the retrospective nature of the case-control study, reverse causation appears in the total cholesterol variable, likely due to post-MI statin use among cases. SHAP analysis captured this treatment effect, highlighting predictive contributions beyond naive associations without implying causality. It also points to the presence of overlooked risk factors that persist despite standard pharmacologic treatment or control. This highlights the need for multidimensional, nonlinear models rather than simple ones. Such approaches are better at capturing the complex interactions underlying multifactorial diseases such as MI.

Still, some limitations should be acknowledged. The dataset represents individuals from a single population, so further research using more diverse cohorts would support broader generalization and build on the performance demonstrated here. As this study used a case–control design, the model’s outputs reflect relationships within the sample and should not be interpreted as absolute risks or incidence rates. Potential confounders may influence some associations. Although SHAP analysis provided valuable insights into feature contributions, it does not establish causal links between periodontal health and MI. Future longitudinal studies are needed to clarify these relationships.

This work contributes from both clinical and conceptual perspectives. From a clinical standpoint, incorporating oral health variables establishes an interdisciplinary framework in which dental professionals can actively participate in the screening and management of cardiovascular risk. Conceptually, the study reframes cardiovascular disease as more than a vascular or metabolic disorder, positioning it instead as a condition influenced by interconnected systems such as immunity, microbial ecology, and behavior. The latter motivates a more systemic understanding of the problem, which constitutes a future extension of our work. By leveraging explainable machine learning, these hidden relationships can be uncovered even in retrospective data, providing not only predictive insight but also a challenge to conventional clinical paradigms.

## Methods

### Study Design and Population

Our model employed data from a population-based case–control study to explore the relation between periodontitis and myocardial infarction^38,39^. Data were collected from two New York counties between 1997 and 2008. Cases (534 subjects) were adult males and females who were hospitalized for a first-time MI and survived. Controls (821 subjects) were drawn at random from the surrounding community to represent the base population.

Comprehensive clinical, behavioral, metabolic, and dental data were collected. Trained dental clinicians conducted periodontal evaluation, including probing pocket depth (PPD) and clinical attachment level (CAL), at multiple sites using standardized periodontal probes. Periodontal health was evaluated based on joint guidelines from the Centers for Disease Control and Prevention (CDC) and the American Academy of Periodontology (AAP)^40^.

The dataset (Table 1) included demographic information, oral health behaviors, like dental visits, and periodontal measurements, such as gingival bleeding and plaque index. Also, lifestyle exposures and cardiometabolic indicators were recorded. All variables were included as potential predictors in subsequent analyses.

Before enrollment, written informed consent was obtained from all participants. All procedures were approved by the institutional review board of the University at Buffalo (IRB #030-488953). Data collection followed ethical standards for research involving human participants, including privacy protections and the voluntary nature of participation.

### Machine Learning Models

A suite of seven supervised machine learning (ML) classifiers was employed to predict myocardial infarction (MI) status using a heterogeneous set of clinical, metabolic, behavioral, and periodontal variables. The classifiers included Logistic Regression (LREG), Random Forest (RF), Extreme Gradient Boosting (XGBoost), K-Nearest Neighbors (KNN), Decision Tree (DT), Support Vector Classifier (SVC), and Naive Bayes (NB). These algorithms were selected to represent diverse modeling paradigms, thus enabling a robust assessment of predictive performance^41^.

Data modeling was conducted using a nested cross-validation framework (see Figure 1), comprising five outer folds for unbiased evaluation and five inner folds for hyperparameter tuning. Within each outer fold, model-specific hyperparameters were optimized via Bayesian optimization, using the area under the receiver operating characteristic curve (AUC) as the objective function. This probabilistic approach models the performance surface as a Gaussian process, allowing for efficient exploration of the hyperparameter space^42^.

Each algorithm was tuned within a custom search space tailored to its properties (see Table 2). The optimal hyperparameters, selected based on average performance across the inner folds, were then used to retrain the model on the full outer training set, which was subsequently evaluated on the corresponding outer test fold^43^.

### Data Preprocessing

The data preprocessing pipeline included two steps: missing value imputation and feature scaling. To handle incomplete data, we utilized the Multivariate Imputation by Chained Equations (MICE) method—an iterative, regression-based technique that models each variable with missing values as a function of the others^44^. This method captures multivariate dependencies and mitigates the bias introduced by simpler imputation strategies. For feature scaling, all continuous variables were normalized to the [0, 1] range using Min-Max scaling, preventing features with larger values from overshadowing others.

To avoid information leakage during preprocessing, both imputation and scaling were exclusively fitted on the training set. The learned transformations were then applied to the test set, ensuring that no information from the test set unfairly influenced model training.

A variant of SMOTE (Synthetic Minority Over-sampling Technique) that supports both numerical and categorical features was applied to address class imbalance^45^. This method generates synthetic samples for the minority class to improve its representation during training. Importantly, oversampling was applied only to the training data to avoid introducing bias into the test results.

### Evaluation Metrics

Model evaluation focused primarily on Area Under the Receiver Operating Characteristic curve (AUC), with supplementary metrics including F1-score and Matthews Correlation Coefficient (MCC) computed to provide a comprehensive performance assessment. Aggregated evaluation results, including cumulative confusion matrices from the outer testing folds, were used to summarize classification behavior across the entire dataset. The definition of the metrics used is as follows:

#### Accuracy

Mainly used with balanced data to measure the proportion of correctly classified samples:

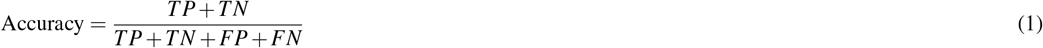

where *TP* denotes true positives, *TN* true negatives, *FP* false positives, and *FN* false negatives.

#### Precision

Shows the proportion of predicted positive cases that are actually positive:

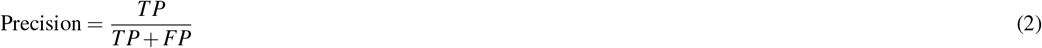

#### Recall (Sensitivity)

Also known as the true positive rate (TPR). It measures the correctly classified proportion of actual positives:

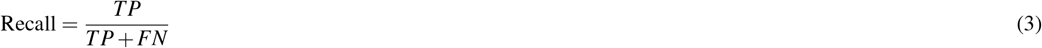

#### F1-Score

It is defined as the harmonic mean of precision and recall, so it symmetrically represents both. It is used when the positive class is the focus.

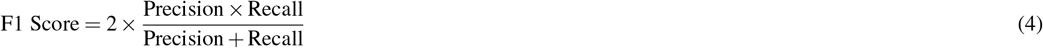

#### Area Under the Receiver Operating Characteristic Curve (AUC)

It represents the probability that a classification model is able to separate the positive from negative classes regardless of the classification threshold. AUC values range from 0 to 1. It is useful for imbalanced datasets.

#### Matthews Correlation Coefficient (MCC)

A robust metric that combines all four parts of the confusion matrix into a single value:

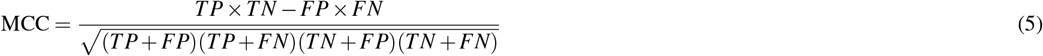

Closely similar to any correlation coefficient, it has a range from ™1 to +1, where +1 indicates perfect agreement and ™1 shows total dissimilarity between predictions and actual labels. Unlike some other metrics that may favor one class, such as accuracy or F1-score, MCC equally includes all classes or parts of the confusion matrix, making it suitable for skewed datasets.

### Calibration Methodology

Well-calibrated probability estimates are especially important in medical applications, where accurate risk predictions help guide clinical decisions and support personalized patient care. For calibration, isotonic regression was applied to the best-performing model (XGBoost) as it is a nonparametric method that makes no assumptions about the shape of the calibration curve and allows flexible adjustment of predicted probabilities. Calibration was performed using the same nested cross-validation framework applied for model evaluation, with isotonic regression fitted only on the training folds to ensure unbiased estimation during testing. To assess calibration visually, reliability diagrams were generated by plotting observed outcome frequencies against predicted probabilities. Calibration curves were obtained separately for each test fold, and the mean calibration curve was calculated and presented. A smoothed 95% confidence interval, estimated via bootstrap resampling across folds, was also plotted to represent fold-to-fold variability, with the bounds smoothed using a Savitzky-Golay filter.

The Brier score was also computed to quantify the calibration quality. It is defined as the mean squared difference between the predicted probabilities and the true binary outcomes:

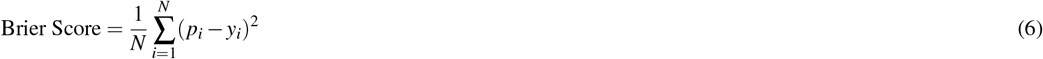

where *p*_*i*_ is the predicted probability for instance *i, y*_*i*_ ∈ {0, 1} is the true outcome, and *N* is the total number of samples. The Brier score ranges from 0 to 1, where lower values indicate better calibration and higher predictive accuracy. A score of 0 represents perfect predictions, while 1 is the worst possible.

### Interpretability Analysis

Following model comparison, SHapley Additive exPlanations (SHAP) were employed to interpret feature contributions in the best-performing model^13^. SHAP values facilitated both global- and individual-level interpretability. At the global level, summary plots illustrate the average magnitude of each variable’s influence on model predictions. At the individual level, waterfall plots highlight the most impactful features contributing to specific patient-level classifications.

## Conclusions

This study shows that periodontitis contributes to systemic health and adds predictive value for myocardial infarction when included in a machine learning model. SHAP analysis shows that periodontal measures like clinical attachment loss, plaque index, and gingival bleeding are strong predictors and reflect systemic inflammation. Our findings suggest that dental data can enhance medical assessment and question the traditional separation of oral and overall health. Including dental features in explainable machine learning models improves prediction and offers clear, personalized insights for medical professionals and patients.

## Acknowledgements

H.H. would like to acknowledge support from the RIG-2023-051 Khalifa University grant. H.H. and P.D. would like to thank the UAE-NIH Collaborative Research grant AJF-NIH-25-KU.

## Author Contributions

Conceptualization and methodology: M.H., H.H.; Data curation: M.H., L.L.; Formal analysis: M.H., H.H.; Resources: P.D., L.L., M.B., K.F.; Writing – original draft: M.H., H.H.; Writing – review and editing: P.D., L.L., K.F., M.B., H.H.; Supervision: H.H.; Funding: P.D., H.H.

All authors approved the final version of the manuscript.

## Data Availability Statement

The dataset utilized in this study is not publicly available but can be obtained from the corresponding author upon reasonable request.

## Additional information

### Competing interests

The authors declare no competing interests.

## Notes

### Competing Interest Statement

The authors have declared no competing interest.

### Author Declarations

Ethics committee/IRB (#030-488953) of the University at Buffalo gave ethical approval for this work

